# Monkeypox in the New York metropolitan area, Summer 2022

**DOI:** 10.1101/2022.11.24.22282177

**Authors:** Rachel Gnanaprakasam, Marina Keller, Rebecca Glassman, Marc Y El Khoury, Donald Chen, Nicholas Feola, Jared Feldman, Vishnu Chaturvedi

## Abstract

Early in the 2022 Monkeypox global outbreak, caseloads in the New York metropolitan area climbed rapidly before other US urban areas. We summarize our monkeypox clinical experience detecting, treating, and vaccinating during a quickly evolving emerging infection.

**Statements:** 

**Ethical statement:** Institutional Review Board exemption was granted for this study.

**Funding statement:** No funding was obtained

**Conflict of interest:** None

**Authors’ contributions:** Each author actively participated in data collection, analysis and editing of the manuscript.

**Collaborators:** None

---

Monkeypox (MPX) is a zoonotic disease caused by a double-stranded DNA monkeypox virus (MPXV) from the *Poxviridae* family that includes smallpox and cowpox viruses [1,2]. Until recently, MPX circulated in a few Western and Central African countries, with cases elsewhere in the globe limited to small, short-lived outbreaks [3,4]. Starting in May of 2022, a global outbreak of monkeypox was reported, which grew to over 77,000 cases in 106 countries [5,6,7]. In the USA, the caseload started to climb rapidly in the New York metropolitan area before outbreak onset in other urban centres of California, Texas, Illinois, and Florida. We summarize here our MPX clinical experience at our 700-bed, level 1 trauma centre, which serves a large swath of the New York metropolitan area, including the lower Hudson Valley and New York City.

The first case of MPX was seen in our hospital in June 2022, in a patient with recent travel to a large social event for gay and bisexual men in Florida. All patients were seen and treated as outpatients either in our specialized MPX isolation clinic or remotely through telemedicine appointments. Many of the patients referred to us were either treated for Human immunodeficiency virus (HIV) infection or were followed for PrEP. The demographics and clinical presentation of 23 MPX patients are summarized in Table 1. Ten patients had a fever, 10 had genital ulcers, 4 had anal ulcers, and 4 had significant proctitis. Two patients were hospitalized for severe proctitis and fever. Ten patients received 11 fourteen-day courses of tecovirimat under an investigational new drug protocol approved by our institutional review board (IRB) within 72 hours of starting the medication. The indication included presence of anal ulcers and proctitis [8,9]. With the initiation of tecovirimat, anorectal pain and bleeding improved within only 3 days in 4 patients. The resolution of one patient’s buttock lesions are shown in Figure 1. The other 6 patients did not notice a marked improvement on therapy but all resolved their symptoms by the end of the 14 days of tecovorimat. No patients noted any side effects attributable to tecovirimat. One HIV positive patient had an especially slow response to his first course of tecovirimat. His initial symptoms recurred several days after stopping the medication. Physical exam revealed new anal ulcers which were swabbed and found to be MPXV positive and Herpes Simplex Virus 1/2 (HSV) negative. A second course of tecovirimat was given which helped resolve his recurrent symptoms. Five patients had received their first dose of Jynneos vaccine within the week prior to symptom onset.

**Table 1.**
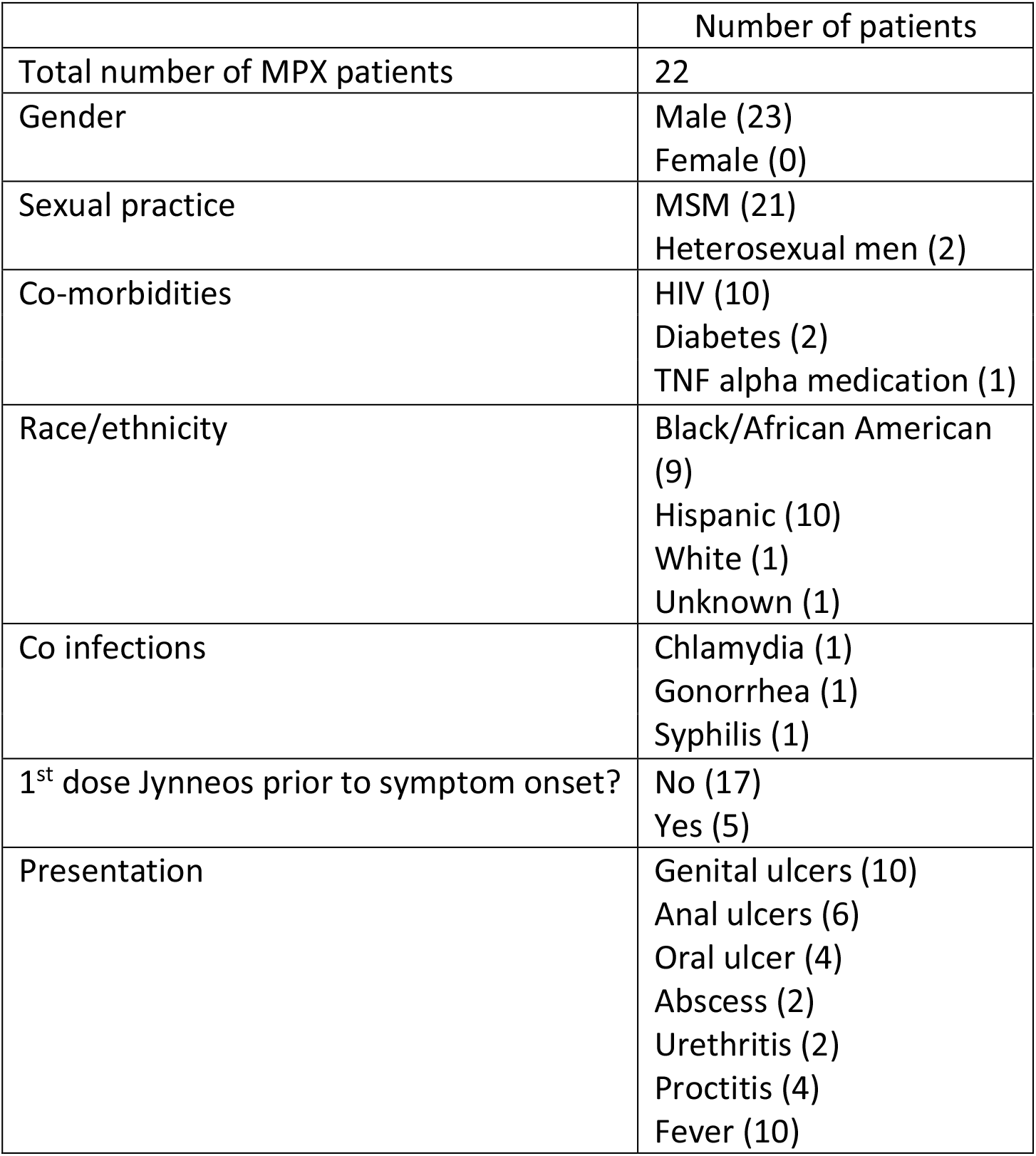
Demographics and clinical outcomes of MPX patients

|  | Number of patients |
| --- | --- |
| Total number of MPX patients | 22 |
| Gender | Male (23)<br>Female (0) |
| Sexual practice | MSM (21)<br>Heterosexual men (2) |
| Co-morbidities | HIV (10)<br>Diabetes (2)<br>TNF alpha medication (1) |
| Race/ethnicity | Black/African American (9)<br>Hispanic (10)<br>White (1)<br>Unknown (1) |
| Co infections | Chlamydia (1)<br>Gonorrhea (1)<br>Syphilis (1) |
| 1<sup>st</sup> dose Jynneos prior to symptom onset? | No (17)<br>Yes (5) |
| Presentation | Genital ulcers (10)<br>Anal ulcers (6)<br>Oral ulcer (4)<br>Abscess (2)<br>Urethritis (2)<br>Proctitis (4)<br>Fever (10) |

**Figure 1.**
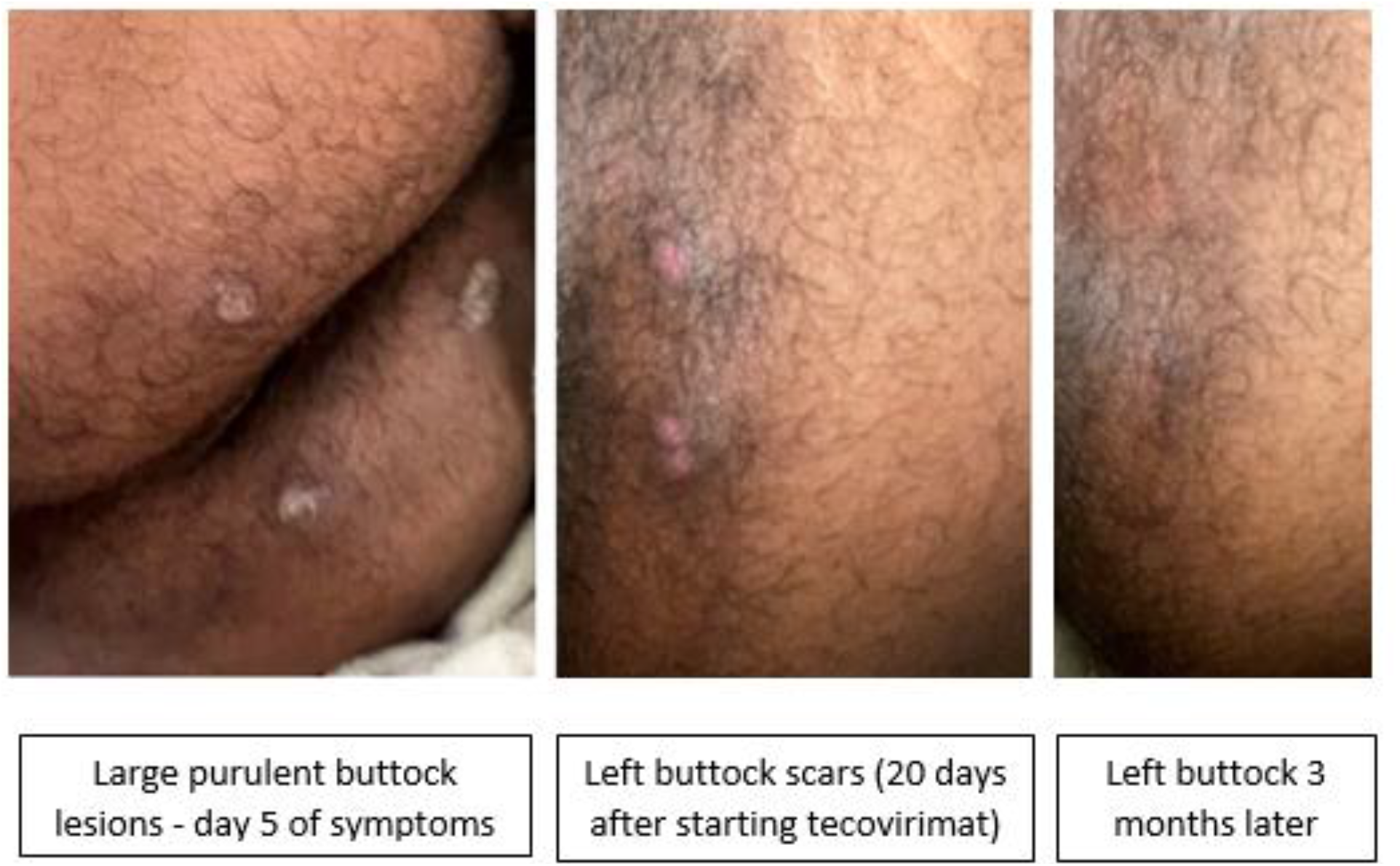

In an effort to curb the spread of MPXV in our community, we collaborated with our county Department of Health and set up a drive thru vaccination centre using the same structure we used for our COVID19 vaccine drive. Our vaccination efforts targeted at-risk populations as defined by the state health department and vaccinated 1,103 people with over 1,800 doses of Jynneos vaccine [10]. The age distribution of vaccinations reflected the age distribution of clinical cases reported in the literature with the highest numbers of vaccinees aged 25-44 (Table 2, Figure 2). Most of our patients (734 people) were New York State residents and 53 patients came from outside the state for the vaccine. There were 107 African Americans, 82 Hispanic and 418 White individuals with a large number of people opting not to give their race or ethnicity (Table 2).

**Table 2.** Demographics of Jynneos vaccine recipients

|  | Number of patients |
| --- | --- |
| Home address | New York State outside New York City (734)<br>New York City (117)<br>Out of state (53)<br>Address unavailable (199) |
| Race/Ethnicity | African American (107)<br>Hispanic (82)<br>White (418)<br>Asian (21)<br>Mixed/other race (70)<br>unknown/refused (405) |
| age | < 24 years old (105)<br>25-44 (547)<br>45-64 (371)<br>>65 (80) |

**Figure 2.**
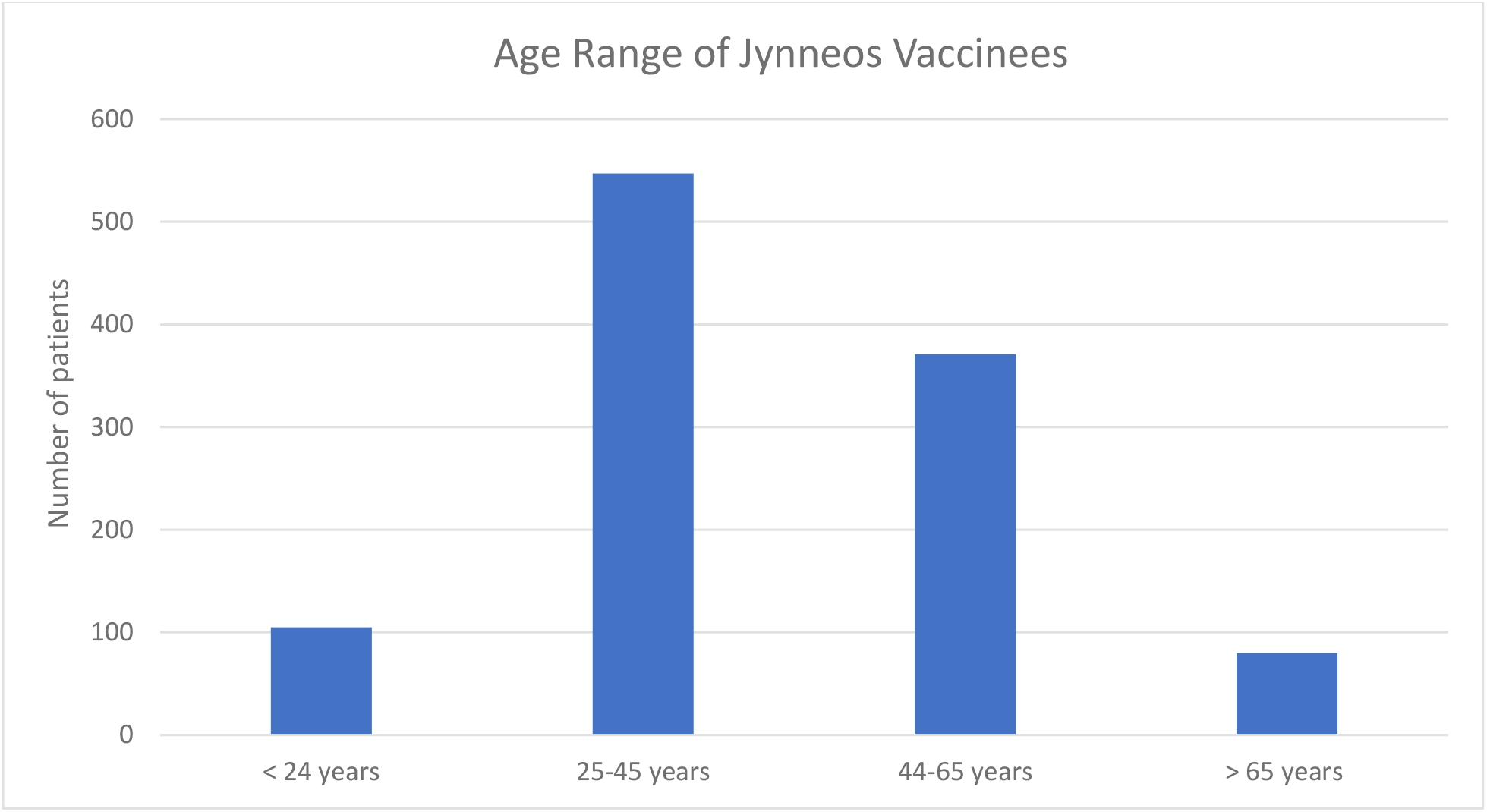

Monkeypox often manifests as a morbid infection eliciting painful mucositis of the pharynx, urethra, and anus. We observed mixed response to tecovirimat with a minority of patients showing rapid clinical response to tecovirimat while the others showed poor to no response. One challenge we faced was the lack of available testing for at-risk patients who presented with symptoms of MPX without cutaneous manifestations due to current restrictions on sample types acceptable for MPXV testing [11]. Similar to the COVID -19 pandemic, we experienced a rapid, high-impact outbreak of MPX in the New York City metropolitan area, requiring urgent mobilization of healthcare resources and close coordination with local and state departments of health. Public health outreach and engagement of at-risk persons, accurate and timely diagnostics, and availability of an investigational drug and active mass vaccination allowed for control of this current Monkeypox outbreak [12].

## Data Availability

All data produced in the present study are available upon reasonable request to the authors

## Acknowledgements

Dr Humayun Islam, Marie Yezzo, Sherita Bernard, Justin Williams, Keri Tone, Dan Amarillo, Diana Guevara, Claudia Dones, Kristy Greene

